# A multilayered post-GWAS analysis pipeline defines functional variants and target genes for systemic lupus erythematosus (SLE)

**DOI:** 10.1101/2023.04.07.23288295

**Authors:** Mehdi Fazel-Najafabadi, Loren L. Looger, Harikrishna Reddy-Rallabandi, Swapan K. Nath

**Author notes:** Correspondence and reprint requests: Swapan K. Nath, PhD, Oklahoma Medical Research Foundation, 825 NE 13^th^ Street, Oklahoma City, OK 73104. Contributed equally (Mehdi Fazel-Najafabadi, Loren L. Looger). Authors declare that the research was conducted in the absence of any commercial or financial relationships that could be construed as a potential conflict of interest.

## Abstract

**Objectives:** Systemic lupus erythematosus (SLE), an autoimmune disease with incompletely understood etiology, has a strong genetic component. Although genome-wide association studies (GWAS) have revealed multiple SLE susceptibility loci and associated single nucleotide polymorphisms (SNPs), the precise causal variants, target genes, cell types, tissues, and mechanisms of action remain largely unknown.

**Methods:** Here, we report a comprehensive post-GWAS analysis using extensive bioinformatics, molecular modeling, and integrative functional genomic and epigenomic analyses to optimize fine-mapping. We compile and cross-reference immune cell-specific expression quantitative trait loci (*cis*- and *trans*-eQTLs) with promoter-capture Hi-C, allele-specific chromatin accessibility, and massively parallel reporter assay data to define predisposing variants and target genes. We experimentally validate a predicted locus using CRISPR/Cas9 genome editing, qPCR, and Western blot.

**Results:** Anchoring on 452 index SNPs, we selected 9,931 high-linkage disequilibrium (r^2^>0.8) SNPs and defined 182 independent non-HLA SLE loci. 3,746 SNPs from 143 loci were identified as regulating 564 unique genes. Target genes are enriched in lupus-related tissues and associated with other autoimmune diseases. Of these, 329 SNPs (106 loci) showed significant allele-specific chromatin accessibility and/or enhancer activity, indicating regulatory potential. Using CRISPR/Cas9, we validated rs57668933 as a functional variant regulating multiple targets, including SLE risk gene *ELF1*, in B-cells.

**Conclusion:** We demonstrate and validate post-GWAS strategies for utilizing multi-dimensional data to prioritize likely causal variants with cognate gene targets underlying SLE pathogenesis. Our results provide a catalog of significantly SLE-associated SNPs and loci, target genes, and likely biochemical mechanisms, to guide experimental characterization.

## INTRODUCTION

Systemic lupus erythematosus (SLE, lupus) is a complex autoimmune disease with substantial genetic underpinnings, *e.g.*, strong familial aggregation(1), large twin concordance (monozygotic>dizygotic)(2), and high sibling recurrence risk ratio (λ_s_∼30)(3). >50 candidate gene studies and genome-wide association studies (GWAS) have identified >100 SLE risk loci (*p*-value<5L×L10^−8^), across multiple ethnicities(4–7). However, these loci explain only ∼30% of SLE heritability (h^2^)(5, 8).

In addition to incomplete knowledge of precise risk loci and alleles underlying GWAS peaks, it is not generally understood how such alleles mechanistically contribute to disease. For a given locus, GWAS often reports a sole (“index”) single nucleotide polymorphism (SNP), which may or may not itself be functional, but is likely in linkage disequilibrium (LD) with disease-predisposing SNPs(9). As in other complex diseases, >90% of reported SLE index SNPs are non-coding (intronic and intergenic). A major challenge in the post-GWAS era is to precisely identify predisposing coding and non-coding SNPs and their associated target genes, and to determine the molecular mechanisms underlying disease risk.

Accurate association determination remains a nontrivial challenge in clinical genomics and genome informatics. Generally, post-GWAS analyses combine multiple GWAS signals using LD structure and epigenetics(10, 11). Additional data sources, such as multiple independent, consistent annotations, greatly assist prioritization of likely functional SNPs(12). SNPs can modulate transcription factor (TF) binding and chromatin structure, altering gene regulation. Indeed, *cis*- and *trans*-expression quantitative trait locus (eQTL) analyses frequently link disease-associated alleles to specific gene/isoform expression(13). Together, annotating open, active chromatin from DNase I hypersensitivity and Assay for Transposase-Accessible Chromatin (ATAC-seq) peaks(14), alongside individual genomic regulatory elements (promoters, enhancers, silencers, *etc*.) using histone marks, chromatin modifiers, and transcription factors, and by *in silico* bioinformatics(15), yields a powerful framework for testing GWAS hypotheses.

Multiple databases (*e.g.*, ENCODE, RoadMap) integrate histone mark data from common cell lines to create consensus regulatory region annotations(16, 17). Moreover, combining ATAC-seq and gene expression data yields chromatin accessibility QTLs (caQTLs) to identify SNPs with allele-specific chromatin effects (*e.g.*, allelic imbalance)(18). Genomic regulatory elements communicate with one another and target genes through complex three-dimensional chromatin interactions (topologically associating domains, TADs). Various chromatin-conformation capture (3C) technologies(19, 20) annotate TADs and other chromatin features. Such interactions, particularly direct enhancer-promoter interactions(21), underlie long-range enhancer activity(22), and can aid GWAS interpretation. Finally, recent methods like Massively Parallel Reporter Assays (MPRAs) simultaneously screen thousands of SNPs for transcriptional enhancer activity, providing information about SNP genomic context and allele behavior(23).

We compiled all available data on the above features and cross-referenced them to predict locus/SNP functionality. Genomic regions can assume different activities in different cell types; we matched datasets taken from the same cell type, ensuring consistency. Further, when possible, annotations were taken from data derived solely from immune cells(15, 21, 24–26), identifying associations relevant to pathogenesis. This approach has revealed target genes and cells associated with rheumatoid arthritis(27) and breast cancer(28), among others. We applied the same regulatory-SNP analysis pipeline to protein-coding SNPs, as many exons contain TF-binding sites and/or promoters/enhancers(29). We also consider the effects of missense SNPs on protein structure/function.

Using an integrated approach, we collate and reassess published SLE-GWAS association signals along with high-LD SNPs, incorporating diverse data on underlying genomic features. For each locus, we define potential functional variants and their cognate target genes in immune cells. We prioritize functional SNPs and assign associated target genes and modulated biochemical pathways. As proof of concept, we utilized CRISPR-Cas9 genome editing, qPCR, and Western blot to validate the allelic effects of a candidate SNP on SLE risk gene *ELF1*.

## MATERIALS AND METHODS

### Study design

Our workflow and study design are shown in **Figure 1**. We 1) collated, from qualified studies, all reported and replicated index and correlated SNPs to define statistically-independent SLE susceptibility loci, 2) predicted SNP effects in statistically-independent loci and annotated them in regulatory tiers, 3) performed molecular modeling on missense SNPs, 4) leveraged cell type-specific *cis-* and –*trans*-expression quantitative trait loci (eQTLs) and promoter-capture Hi-C (PCHiC) data to define “enhancer” and “promoter” SNPs and target genes, 5) estimated locus overrepresentation in molecular pathways and gene ontology categories and identified cell type-specific SNP enrichment in epigenetic features, 6) used chromatin accessibility QTLs (caQTLs) and massively parallel reporter assay (MPRA) data to identify allele-specific effects, and 7) experimentally validated a functional variant using CRISPR/Cas9- based activation/silencing in B-cells.

**Figure 1.**
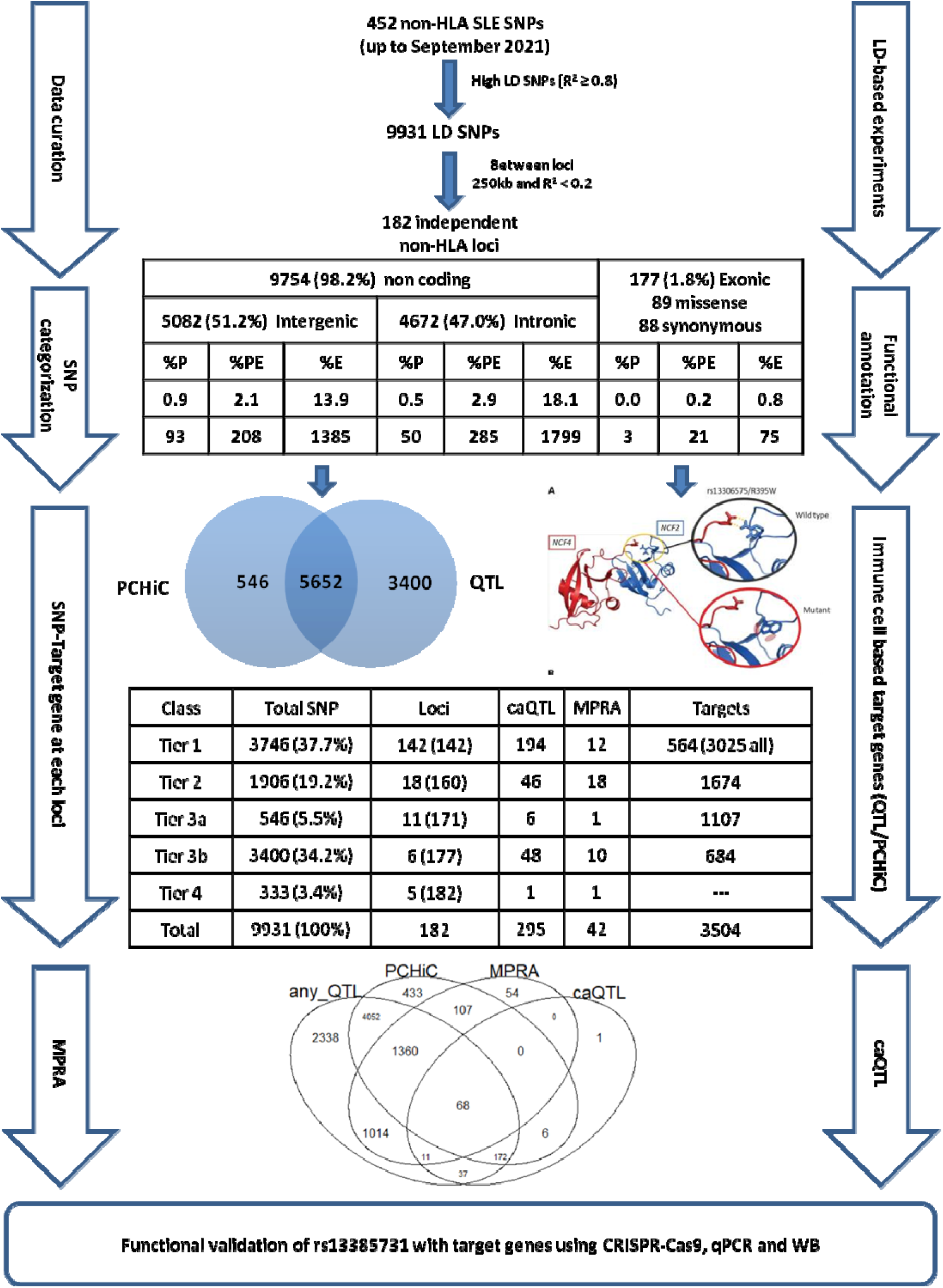
Study framework and summary. Tier 1: PCHiC and QTL with at least one same target gene, Tier 2: PCHiC and QTL with different targets, Tier 3a: Only PCHiC target, Tier 3b: Only QTL target, Tier 4: No targets.

**Figure 2.**
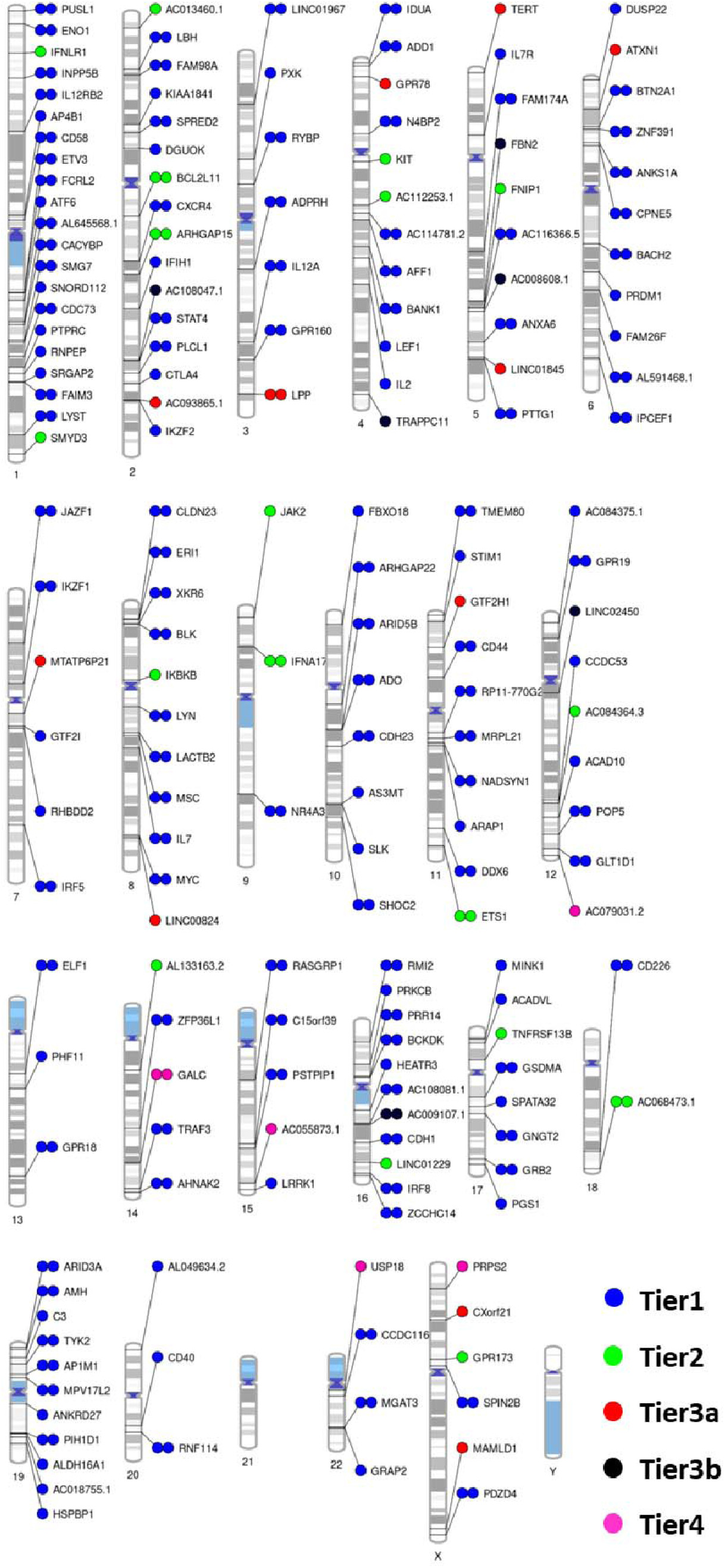
Distribution of 182 non-HLA SLE loci across th human genome. Loci colored by highest SNP tier. Tier1 names are common target genes from both eQTL and PCHiC data; other tiers are named by the closest positional gene. Loci with double dot have ≥1 significant experimentally validated (caQTL or MPRA) allele-specific SNP. Single dots mean no experimentally validated SNPs are yet known.

### Collating variants

We thoroughly reviewed SLE association studies up to September 2021, encompassing GWAS and candidate-gene studies with sample sizes >2,000. We selected genome-wide significant index SNPs (P<5x10-8) (**Supplementary Table 1, Supplementary Notes**). To find potential causal SNPs, we expanded each locus to its LD region, treating loci as independent if separated by >250 kb with low LD (r2<0.2). We excluded the HLA region.

### Regulatory region annotation

We employed diverse bioinformatics tools and databases to assess the regulatory implications of each SNP (**Supplementary Notes**). To identify allele-specific enhancers, we conducted Massively Parallel Reporter Assays (MPRAs) involving over 3,000 SNPs with both alleles present(30). Furthermore, we utilized chromatin accessibility quantitative trait loci (caQTLs)(18, 24) data to enhance the fine-mapping and annotation of SNP-specific regulatory elements.

### Target genes

We used two methods to determine SNP targets **(Supplementary Notes)**. First, SNPs were annotated with *cis*- and/or *trans-*expression QTLs (eQTLs) and splicing QTLs (sQTLs) using multiple databases. Second, to identify SNPs interacting with enhancers and promoters through chromatin interactions, we overlapped associated SNPs within anchors of chromatin interactions in immune cells with available promoter-capture Hi-C (PCHiC)(15, 21) data from immune cells.

### SNP/geneset enrichment analysis

Gene targets of functional SNPs were tested for enrichment in Gene Ontology (GO) categories, biochemical pathway membership, and disease association. Enrichment analysis was carried out using FUMA(11) and epiCOLOC(31) on different SNP sets and their target genes identified through target-type annotations.

### Transcription factor binding

Binding sites were annotated from UCSC Genome Browser GRCh37/hg19 JASPAR core 22.

### Protein models

Protein models were taken from AlphaFold2 and illustrated with PyMOL.

### Cell culture and transfection

Lymphoblastoid cell lines (LCLs; NA18566) with the TT genotype were thawed and cultured in T25 culture flasks until they reached a confluence of 0.5 - 0.7 x 10^6 cells/mL. For CRISPR-based inhibition and activation, we used the plasmids SP- dCas9-TET1 and SP-dCas9-LSD1. We co-transfected pools of *ELF1*-sgRNA plasmids with dCas9-based activation and inhibition plasmids into LCL cells using electroporation. Cells transfected only with sgRNA plasmids were used as the control group.

### CRISPR-based functional validation

We used CRISPR/Cas9 activation/silencing (CRISPRa/i) to bring activating or silencing domains to rs57668933. Briefly, single-guide RNA (sgRNA)/Cas9-RNP complex was prepared at room temperature in Cas9 buffer. RNP complex was transfected into NA18535 LCL cells with electroporation and allowed to express for 72 hours.

### qPCR

To assess the impact of the SNP on target gene expression, we used qRT-PCR on WT, CRISPRa, and CRISPRi cells, as described elsewhere(32). RNA was isolated from WT and CRISPRa/i cells using an RNA Mini kit (Zymo Research) and reverse transcribed using iScript Reverse Transcription Supermix cDNA synthesis kit (Bio-Rad). We measured *ELF1* expression and analyzed results for significance using Prism V.7 (GraphPad).

### Western blot

Cells were collected 72 hours after transfection and lysed in RIPA buffer supplemented with a protease and phosphatase inhibitor cocktail (as outlined in **Supplementary Notes**). The blot was visualized using an Azure ChemiBlot machine, and the obtained results were subjected to analysis. Expression levels were quantified using ImageJ, and densitometry values were graphed using GraphPad Prism.

## RESULTS

### Defining independent SLE candidate loci

Overall, we identified 452 reported genome-wide significant (P<5x10^-8^) non-HLA index SNPs from 76 different GWAS and candidate gene studies (**Supplementary Table 1**). Most index SNPs derived from East Asian and European ancestry studies (**Supplementary Figure 1**). Most (242, 53.5%) index SNPs lay within 145 genes, 210 (46.5%) were intergenic (**Supplementary Table 2**).

We then collected SNPs in high (r^2^>0.8) linkage disequilibrium (LD) with index SNPs, finding 9,479 – totaling 9,931 SNPs for study. We binned these into 182 statistically independent loci (**Table 1, Supplementary Table 2**), with median locus size of 57.7 kb [range 314 bp – 1.15 Mb]. Of the 182 loci, 89 contained single index SNPs; the rest had 2-14 (median 2; **Supplementary Figure 1, Supplementary Table 7**). Total linked SNPs per locus ranged from 1-1,148 (median 26; **Supplementary Table 7**). Correlated SNPs per index SNP ranged from 1-146 (median 24). Fifteen loci had single index SNPs and no LD-SNPs; conversely, LOC_180 had two index SNPs and 1,146 LD-SNPs. The physical distance between index SNPs and LD-SNPs varied from 1 bp to 499 kb (median 14 kb).

**Table 1.**
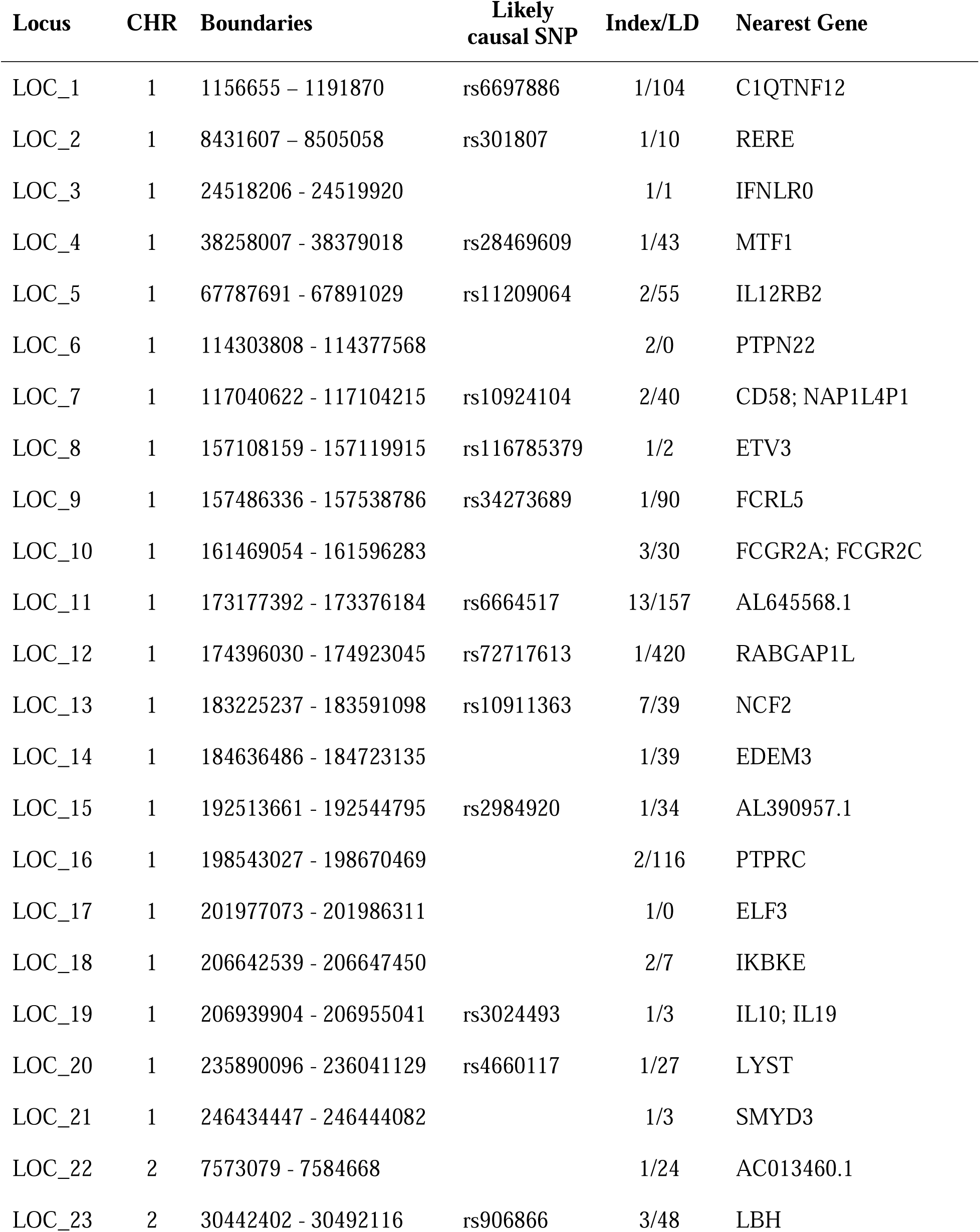
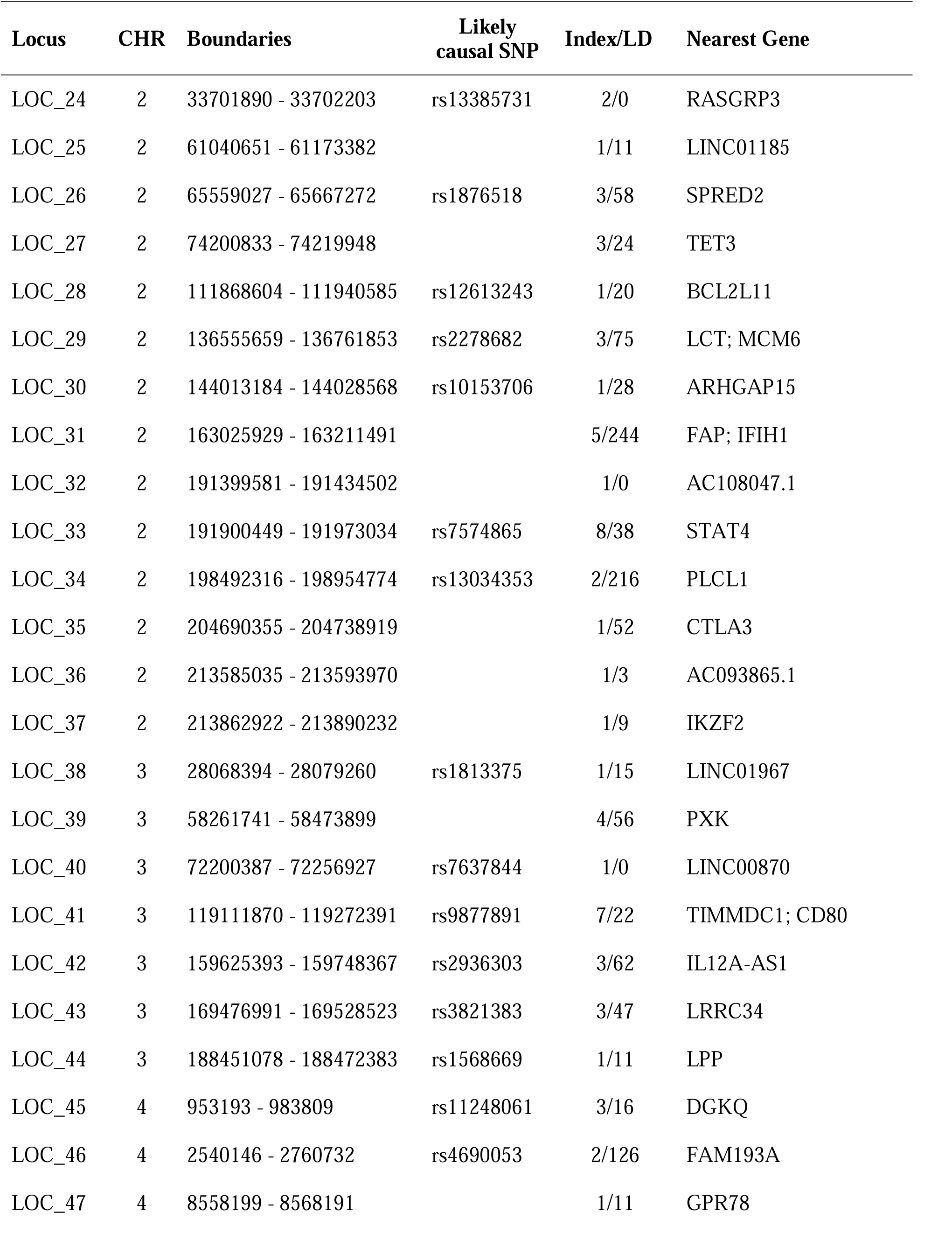
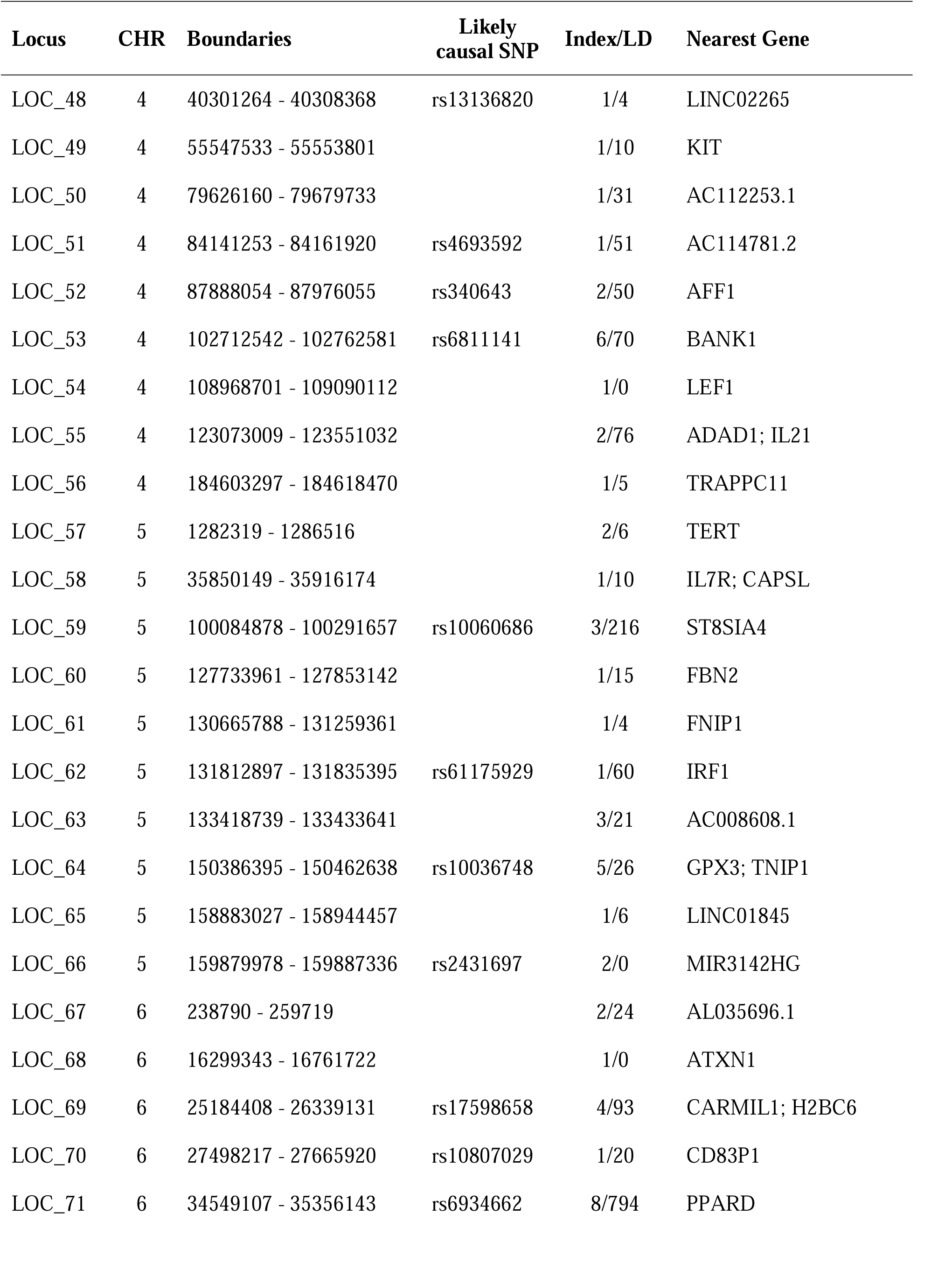
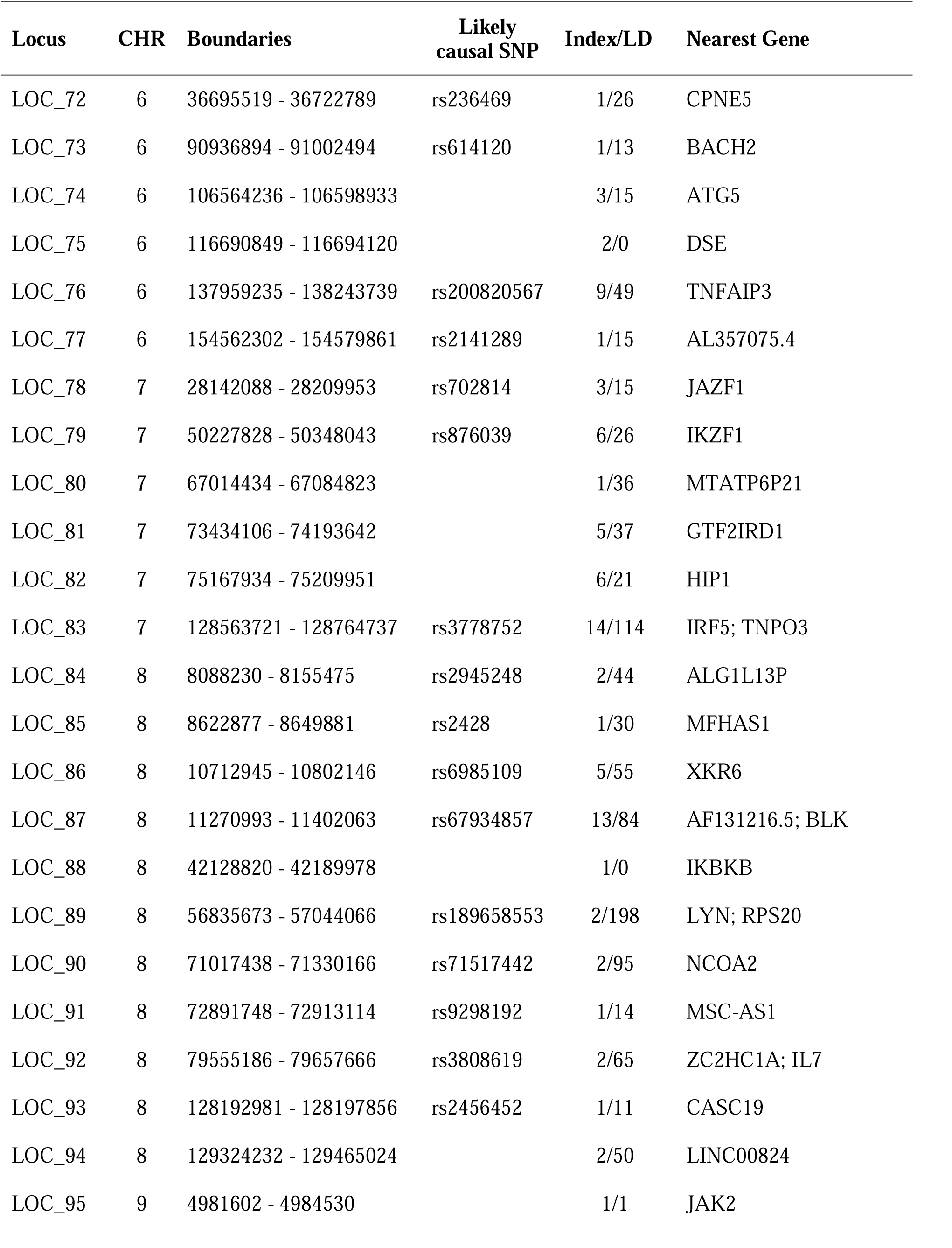
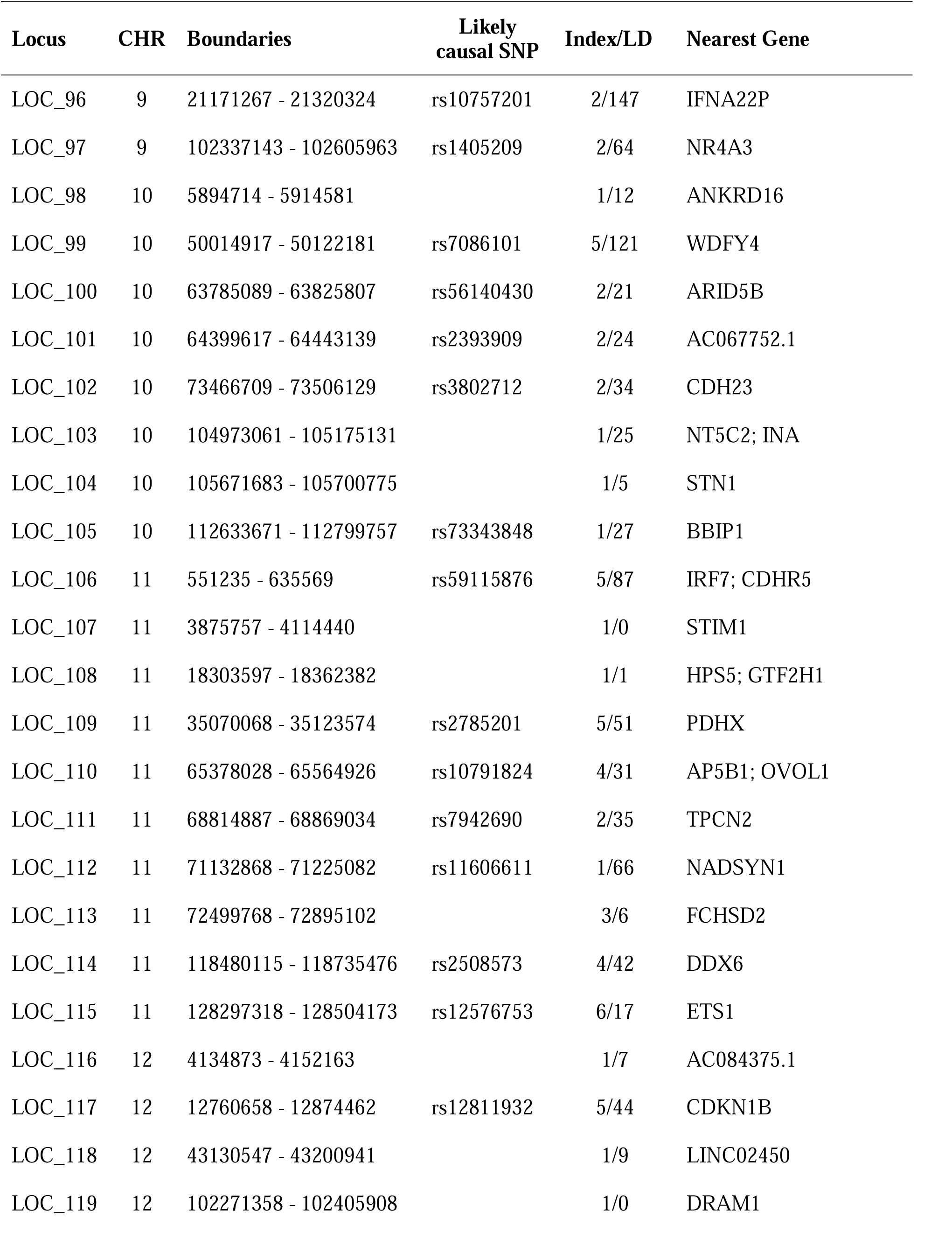
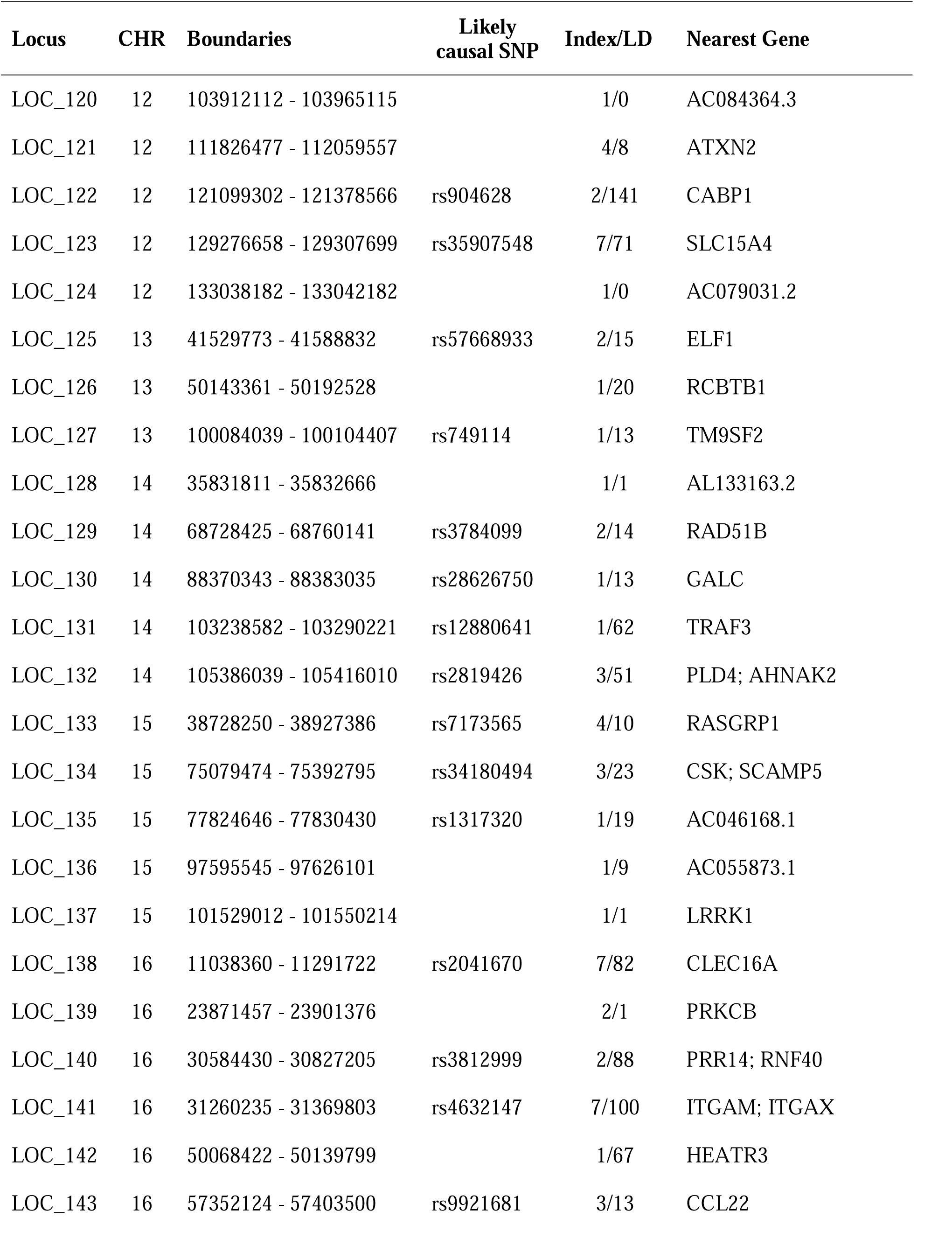
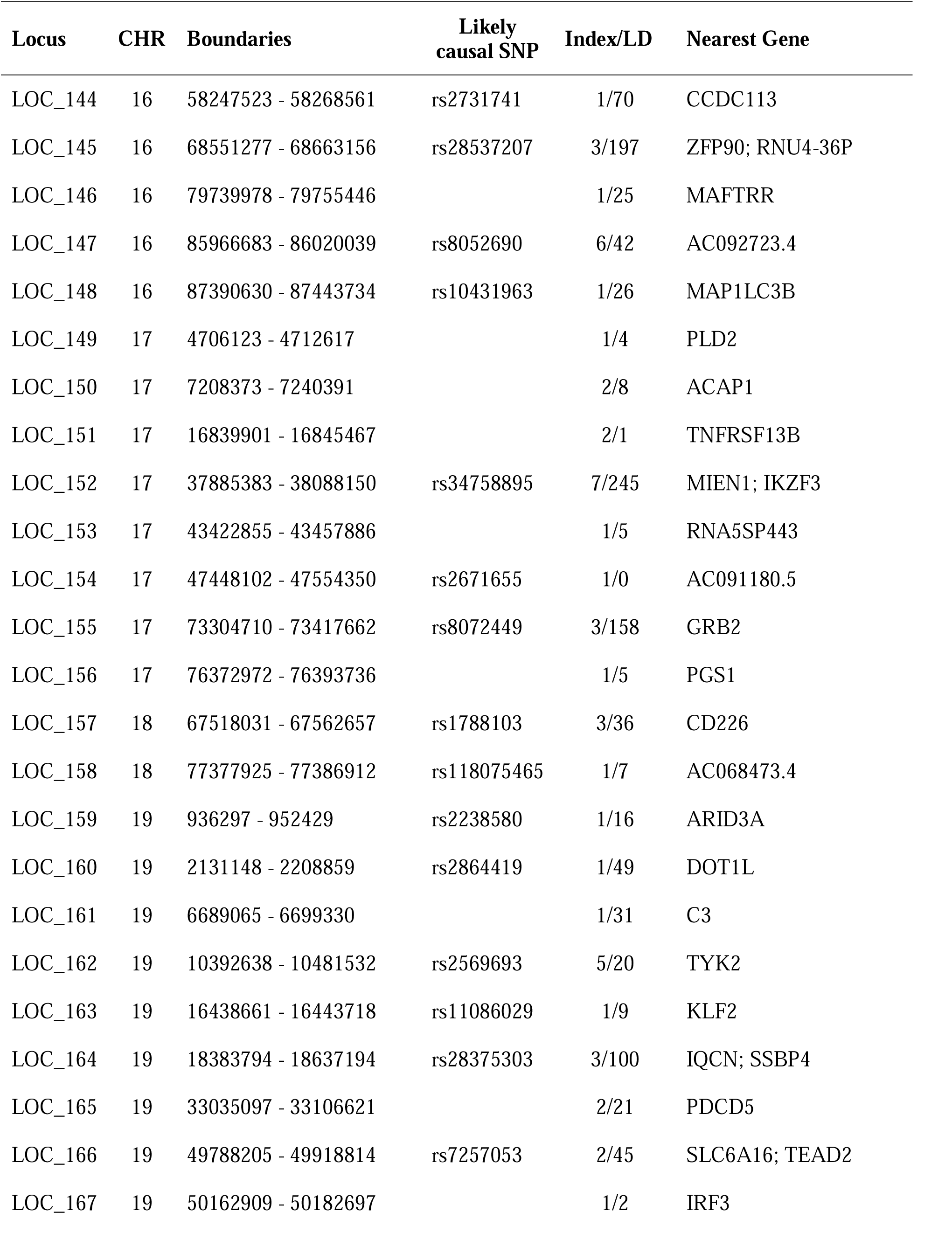
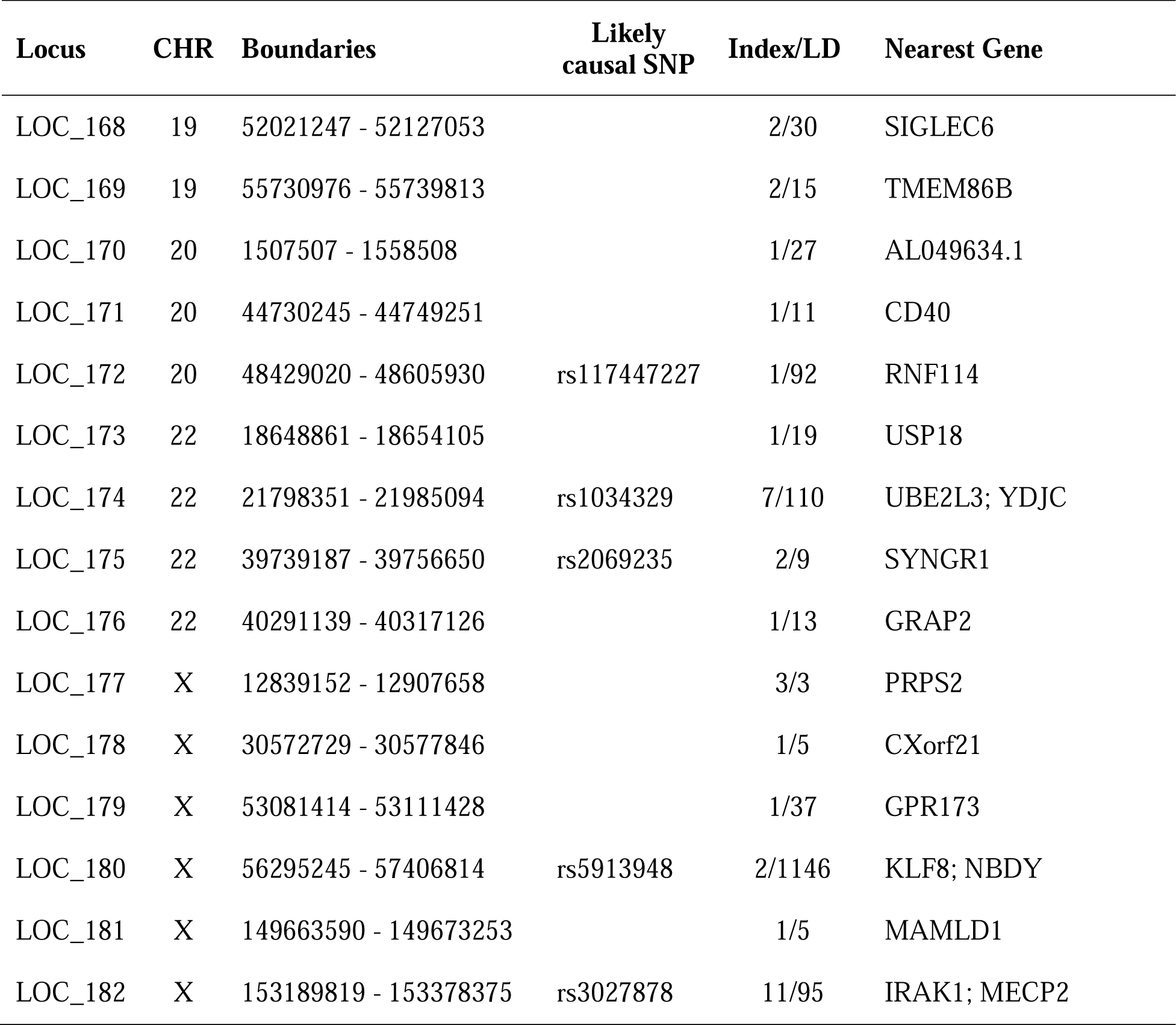
Summary of all SLE loci.

The 182 independent loci contain 426 genes. Of 9,931 total SNPs, 47.0% are intronic, 0.9% synonymous, 0.9% missense, and 51.2% intergenic (**Figure 1**, **Supplementary Table 2**). We annotated all SNPs for *cis*- and *trans*-regulatory effects. The 89 missense SNPs also potentially alter protein structure/function/expression and were molecularly modeled.

### Annotation pipeline

To annotate and prioritize these 9,931 SNPs at 182 loci/426 genes, we established this pipeline: 1) collate eQTLs and 2) PCHiC from immune cells (**Supplementary Table 6**), 3) combine to initially classify SNPs, 4) add histone mark and MPRA data, 5) refine GWAS peaks with caQTLs, 6) experimentally test prioritized SNPs. caQTLs appear much narrower than many other GWAS signals(18); however, of immune cells, they are currently only available for B-cells. As such, we placed them late in our pipeline, so that the initial prioritization covers all cell types. As caQTL data becomes more widely available for other cell types, placing this step earlier in the pipeline could narrow GWAS peaks sooner.

### eQTLs

We first annotated all SNPs with *cis-* and *trans*-eQTLs and associated target genes, using only immune cell-specific data. Most SNPs (9,052) have ≥1 significant *cis*-eQTL [range 0-31; 856 SNPs have single *cis*-eQTLs and 5,539 have ≤5 *cis*-eQTLs] (**Supplementary Table 2**). *cis*-eQTL targets are enriched in immune-related genes, with many being known SLE risk loci. In LOC_13, rs17849501 (Neutrophil cytosol factor 2, *NCF2*) is an eQTL of several genes in multiple immune cell types. We experimentally demonstrated strong, allele-dependent enhancer activity of this SNP(33). In LOC_66, rs2431697 (intergenic) affects expression of multiple genes across cell types. This SNP has been experimentally shown to physically associate with the promoter of miRNA-146a, a potent immune regulator(34) and SLE biomarker(35). In LOC_76, SLE risk SNP rs2230926 (Tumor necrosis factor, alpha-induced protein 3, *TNFAIP3*) greatly increases neutrophil extracellular traps and citrullinated epitopes in SLE patients(36). In LOC_83, rs13239597 (intergenic) is an experimentally validated allele-specific enhancer of Interferon regulatory factor 5 (*IRF5*), a key SLE risk gene.

For *trans*-eQTLs (having target genes >1 Mb or on another chromosome), we identified 75 SNPs from 22 loci targeting 272 unique genes (range 1-149 per locus; 20 out of 22 *trans*-eQTLs had 1-11 target genes) with false-discovery rate (FDR) <1e-5 (**Supplementary Tables 8-10**). Among them, 13 target genes were distal, and 259 on different chromosomes. Among 75 *trans*-eQTL SNPs, 73 were also identified as *cis-*eQTLs. At LOC_121 (*SH2B3*, *ATXN2*), all 8 *trans*-eQTL SNPs showed >100 target genes, demonstrating substantial interactions across the genome. *SH2B3* (*a.k.a.* lymphocyte adaptor protein, *LNK*) links numerous immune signaling pathways to inflammation(37) and is a major immune regulator. LOC_79 (*IKZF1*) had one *trans*-eQTL (rs4917014) with 50 target genes. Many target genes were themselves immune-related and often SLE-associated. rs1990760, a coding SNP at *IFIH1* (LOC_31), is defined for lupus susceptibility(38). This SNP is also a *trans*-eQTL targeting nine genes (*MX1, IFI44L/IFI44, HERC5, IFIT1/IFI6, OAS3/OAS2, HERC6*) significantly enriched in type I/II interferon signaling genes. Interestingly, seven (*MX1, IFI44L/IFI44, HERC5, IFIT1/IFI6, OAS3*) and four (*MX1, IFI44L/IFI44*, *HERC5)* target genes were also targeted by LOC_97 and LOC_99, respectively (**Supplementary Table 8**), suggesting that coregulation of core genes further amplifies *trans*- effects in an omnigenic model(39).

### Chromatin interactions

We independently analyzed PCHiC data on immune cells(15, 21). The 6,198 SNPs had ≥1 PCHiC connection (762 SNPs had 1; 3,322 SNPs had ≤5; maximum 93). Combining eQTL and PCHiC datasets, our SNPs target 3,504 unique genes (**Figure 1, Supplementary Tables 2, 4**).

### SNP categorization

Concordance between eQTL and PCHiC annotation suggests that a given SNP has a strong regulatory role; thus, we based SNP tiers on this intersection (**Figure 1, Supplementary Table 2**). Tier1 includes SNPs annotated by both methods with non-zero target gene overlap. These SNPs (3,746 from 143 loci) have strong evidence of controlling expression of specific target genes. The 1,906 SNPs (17 loci; Tier2) were annotated by both methods but targeted different genes in existing datasets. Tiers 3a and 3b (546 SNPs, 11 loci; 3,400 SNPs, 6 loci) showed either PCHiC or eQTL activity, respectively, but not both. Finally, 333 (Tier4) exhibited neither activity. Of 9,052 *cis*-eQTL SNPs, 3,746, 1,906, and 3,400 were categorized as Tier1, Tier2, and Tier3b, respectively. Of 75 *trans*-eQTL SNPs, 50, 11, and 14 were Tier1, Tier2, and Tier3b, respectively.

### Regulatory elements

Linked SNPs were closely associated with transcriptional regulatory regions annotated by GenoSTAN and other databases; 4,332 (43.6%) lie in annotated promoter, enhancer, and/or silencer regions (**Supplementary Table 2**). Of 9,059 eQTL SNPs, 3,457 (38.1%) lie in enhancers, 625 (6.9%) in promoters, 485 (5.4%) in both, and 670 (7.3%) in silencers. We observed median 13 transcriptional element-associated SNPs per locus (4 loci had no such SNPs; LOC_71 had 360). The bulk were Tier1/Tier2 SNPs, indicating a relationship between transcriptional regulatory elements and eQTL/PCHiC activity. Enhancer SNPs that are also eQTL SNPs had a median distance of 47.2 kb to their target genes’ transcription start sites (TSSs); for Tier1 SNPs, this distance was 45.0 kb. Enhancer SNPs that are also PCHiC SNPs had a median distance of 214.5 kb to their target genes’ TSS; for Tier1 SNPs, this distance was 193.3 kb (**Supplementary Table 4)**. Tier1 SNPs are substantially closer to their target genes than other tiers, consistent with stronger regulatory effects.

Of all regulatory element-associated SNPs, 117 (from 32 loci) were Tier1 SNPs with 1-4 common target genes, leading to 58 unique genes targeted in both eQTLs and PCHiC. Of enhancer SNPs, 93 (26 loci) were Tier1, together targeting 44 unique genes (**Supplementary Table 2**). These SNPs, which are in annotated enhancers, are involved in chromatin interactions, and transcriptionally regulate specific target genes, represent highly prioritized candidates and are given further attention below.

### Massively parallel reporter assays

As an independent measure of SNP effects on transcription, we mined massively parallel reporter assay (MPRA) datasets, which characterize enhancers in high-throughput(40). We examined MPRA data from B-cells (GM12878)(30). A total of 2,614 SNPs appeared in this dataset, and 42 out of 51 significant ones showed allele-specific expression (ASE; FDR<0.01; **Supplementary Tables 12, 13**). MPRA-ASE SNPs were overwhelmingly non-coding: 50 intergenic, 46 intronic, 1 synonymous, 1 missense.

### Deleteriousness scores

We annotated SNPs with pre-computed deleteriousness scores (predictSNP2, CADD, GWAVA). Of exonic SNPs (177 in 61 loci, 91 unique protein-coding genes; 89 missense from 43 loci, 57 unique genes), the algorithms identified 11, 26, and 37 deleterious SNPs, respectively. For missense SNPs, 9, 17, and 37, respectively, were labeled deleterious. For non-coding SNPs, 516 (55% intronic, 45% intergenic) were deemed deleterious by ≥1 algorithm (**Supplementary Table 3**).

### Chromatin accessibility

We next annotated our SNPs according to two measures of chromatin accessibility in whole blood: DNase hypersensitivity and ATAC-seq. Tier1 had by far the largest signals, followed by Tier2 and 3a (**Supplementary Figure 4**). Tiers 3b and 4 showed essentially zero enrichment.

### caQTL SNPs

To identify SNPs with allele-specific chromatin accessibility, we searched a caQTL database from lymphoblastoid (B-cell) cell lines (LCLs) from ten ethnicities(18). caQTL peaks are quite narrow(41); however, the method is new and of immune cells, has only yet been applied to LCLs. Thus, although the technique dramatically reduces SNP numbers, the results here are specific to B-cells. SLE, of course, manifests through numerous cell types; this analysis is only a subset of associated SNPs. As caQTLs are determined in more cell types, this analysis can be extended.

Of our SNPs (covering 100 loci), 295 are caQTLs in ≥1 ancestry. Among our 182 loci, 100 had ≥1 caQTL SNP (range 1-16); 73 loci had ≥1 Tier1 caQTL SNP (**Figure 1**). All but one caQTL SNP were also eQTL SNPs. Of 295 caQTL SNPs, 194 are Tier1, 46 Tier2, 6 Tier3a, 48 Tier3b, and 1 Tier4. Thus, caQTL SNPs are heavily enriched in high-tier SNPs, illustrating that unsurprisingly, SNP-driven changes in chromatin accessibility strongly contribute to downstream expression and chromatin interaction phenotypes. Of 295 caQTL SNPs, 235 (79.7%) lie in enhancers, 91 (30.8%) in promoters, 63 (21.4%) in both, and 19 (6.4%) in silencers. This is consistent with eQTL and MPRA data, although caQTL SNPs are substantially more enriched in enhancer SNPs (**Supplementary Table 11**).

### Transcription factor binding

Next, we independently annotated transcription factor (TF) binding sites using epiCOLOC(31). Tier1 SNPs showed by far the most TFs (89) with binding site enrichment (**Supplementary Figure 3**), with Tier2 next. Tier3a showed small enrichment, and Tiers 3b and 4 were negligible. TFs highly represented in Tier1/Tier2 SNPs include Brachyury/TBXT, TCF4, MYB, and NFKB1–all critical immune-linked proteins involved in SLE pathogenesis. Altogether, TFBS enrichment strongly correlates with eQTL/PCHiC activity, and enriched TFs were immune-linked and SLE-associated.

### Tissue enrichment

Next, for our collected loci, we tabulated expression in diverse tissues using FUMA GENE2FUNC. Tier1 loci target genes were significantly enriched (FDR <0.001) in whole blood and lymphocytes (**Supplementary Figure 2**). As before, lower tiers were much less enriched in these tissues and demonstrated less tissue enrichment overall.

### Disease and pathway association

We searched disease GWAS association catalogs; Tier1 loci target genes were significantly overrepresented in 154 out of 310 traits/diseases. SLE, rheumatoid arthritis (RA), and inflammatory bowel disease (IBD) were particularly enriched in GWAS hitting these loci. Lower tiers were much less linked to disease GWAS. Similarly, Tier1 loci target genes were highly enriched among KEGG pathways (36 out of 68) and gene ontology (GO) classifications (464 out of 1,374), whereas lower tiers were not. Tier1 loci-associated pathways included immune system regulation, cytokine production, phosphorus metabolism, and regulation of protein modification and interferon signaling (**Supplementary Table 5)**. Further studies are required to flesh out exact pathways and mechanisms by which these highly associated SNPs contribute to dyshomeostasis and SLE progression; these results will prioritize avenues for experimental investigation.

### Missense SNPs

We highlight several missense SNPs predicted to dramatically disrupt protein function. rs78555129 mutates a universally conserved arginine in adipolin (CTRP12/FAM132A/C1QTNF12) to cysteine, perturbing protein folding and presumably interactions with its (currently unknown) receptor (**Supplementary Figure 5a**). Adipolin is an anti-inflammatory adipokine implicated in diabetes, arthritis, and obesity(42). In B-cell scaffold protein with ankyrin repeats 1 (BANK1), rs10516487 destabilizes the protein (**Supplementary Figure 5b**), likely interfering with its interactions with TRAF6 and MyD88 in innate immune signaling(43). rs201802880 in Neutrophil Cytosolic Factor 1 (NCF1/p47phox) mutates a universally conserved residue (**Supplementary Figure 5c**), leading to protein destabilization. NCF1 is a subunit of NADPH oxidase, critical for phagocytic immune responses(44). rs2230926 in TNFα-Induced Protein 3 (TNFAIP3) mutates a universally conserved residue important for protein stability (**Supplementary Figure 5d**). TNFAIP3 is indispensable to TNF signaling and immune activation and is an SLE risk gene(45).

### CRISPR-based validation of rs57668933

To validate our approach, we employed CRISPR activation (CRISPRa) and inhibition (CRISPRi) targeting the rs57668933 locus (**Figure 3e**). Both activation domains doubled *ELF1* transcript levels, while both suppressor domains halved them, as confirmed by Western blot (**Figure 3f**). The CRISPR-dCas9-based activation and inhibition system revealed distinct alterations in ELF1 protein expression. Compared to the control group transfected with sgRNA only, the dCas9-TET1 activation plasmid significantly increased (∼1.8x) ELF1 protein expression, while the dCas9-LSD1 inhibition system reduced (∼0.8x) ELF1 expression. These results strongly support the notion that the SNP region plays a pivotal role in regulating *ELF1* expression, consistent with our other findings.

**Figure 3.**
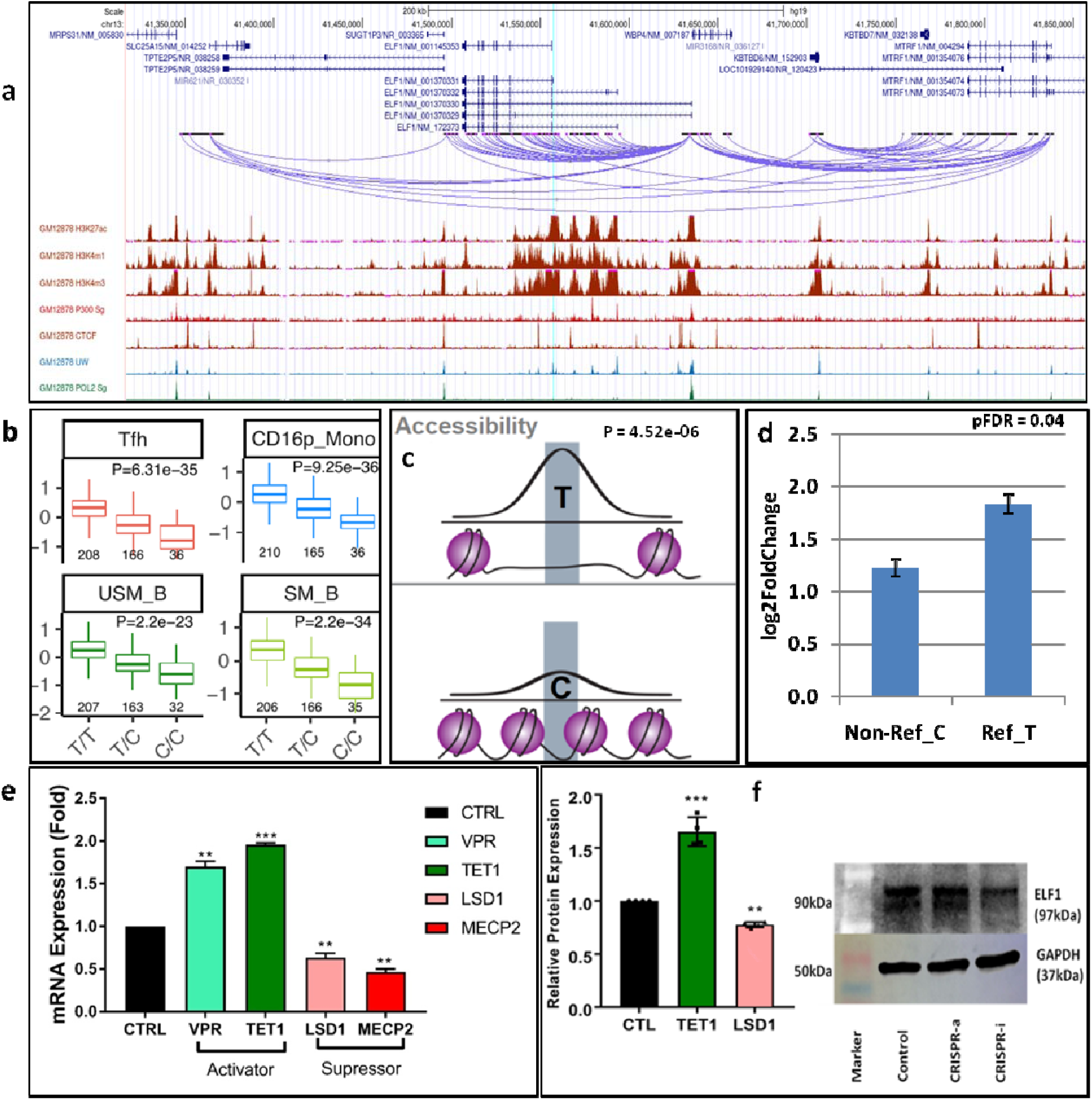
Analysis of LOC_125. This locus has two index SNPs and 15 high-LD SNPs. Of these, rs57668933 is a caQTL-SNP with allele-specific enhancer activity. **a)** Visualized connections of rs57668933 and neighboring regions based on PCHiC, alongside various histon marks. **b)** The SNP lies in *ELF1*, a significant eQTL gene. Significant genotype-specific gene expression in follicular T-helper cells (Tfh), CD16^+^ monocytes, and unstimulated and stimulated B-cells. **c)** Chromatin accessibility of alleles. Reference allele T has higher chromatin accessibility. **d)** MPRA data shows T has higher enhancer activity. **e)** CRISPR-dCas9-based targeting of rs57668933-containing region by two activators (dCas9-VPR and dCas9-TET1) and two suppressors (dCas9-MECP2, dCas9-LSD1) and *ELF1* (target gene) mRNA expression. **f)** Western blot demonstrating the differential expression of ELF1 protein in response to the CRISPR-dCas9-based activation (TET1) and inhibition (LSD1) systems. Densitometry plot illustrating representative Western blot results for ELF1 protein expression. Higher expression is observed in the presence of dCas9-TET1 (activator, lane 2), while reduced expression is observed in the presence of dCas9-LSD1 (inhibitor, lane 3) compared to the control (lane 1). Significance values (**p < 0.005, ***p<0.0005) indicate statistically significant differences.

## DISCUSSION

We have established a state-of-the-art SNP and locus analysis pipeline for assimilating data regarding gene expression, chromatin accessibility and interactions, histone marks, transcription factor binding, tissue expression, and disease association. Our pipeline dramatically reduces large sets of associated SNPs to several likely causal SNPs for experimental validation. This pipeline will be useful for diverse genetic association studies.

After carefully gathering all high-quality SLE GWAS and candidate gene studies up to September 2021 and their high-LD SNPs from 1000Genomes Project Phase3, we defined 182 statistically independent, non-HLA loci totaling ∼10,000 SNPs. Our analysis first focused on SNPs with effects on gene expression; unsurprisingly, these SNPs were overwhelmingly non-coding, and very often localized to enhancer regions. We also found many missense SLE-associated SNPs. These SNPs had high deleteriousness scores; in fact, 30% of the most deleterious SNPs were missense, compared to 1.7% of all SNPs. This dramatic enrichment supports their involvement; it should be noted, though, that CADD and other programs generally view missense SNPs as fairly deleterious. In further support, molecular modeling showed that many missense SNPs adversely affect protein structure and function.

Intriguingly, we found several examples of SNPs encompassing both effects: they were simultaneously missense SNPs with adverse predicted effects on protein function, and enhancer SNPs affecting expression of multiple other genes. For instance, we again found rs1143679, which mutates a key protein residue of integrin alpha M (ITGAM) and disrupts multiple transcription factor binding sites, dramatically weakening enhancer activity(46).

Only 45 loci contained missense SNPs; most SNPs were non-coding. Our pipeline tiered SNPs according to target gene expression (eQTL) and chromatin interactions (PCHiC). We obtained 3,746 Tier1 SNPs, where the two independent experiments identified common regulated genes. Of these, 1,913 are also enhancer-SNPs. Overall, 100 loci had ≥1 caQTL SNP (total 295), and 22 loci had ≥1 allele-specific enhancer SNP (total 42). Together, these constitute 106 out of 182 total SLE loci (329 total SNPs) flagged by ≥3 independent experimental methods regarding gene regulation: eQTL-chromatin interaction-chromatin accessibility (B-cells) or eQTL-chromatin interaction-enhancer histone marks (**Table 1, Supplementary Table 2**). Adding MPRA data (GM12878 cells), yielded a final set of six loci (6 SNPs; **Table 2**) flagged by all available experimental methods with highly significant changes in eQTLs, chromatin accessibility, and target gene expression. These SNPs are predicted to be highly associated with SLE, with effects manifested through enhancer-driven alteration of target gene expression, mediated through B-cells.

**Table 2.** Six most significant SNPs and their target genes.

| rsID | Chr | Risk/Non-Risk | Closest Gene | Common target genes |
| --- | --- | --- | --- | --- |
| rs13385731 | 2 | T/C | *RASGRP3* | *RASGRP3, FAM98A* |
| rs2936303 | 3 | G/A | *IL12A-AS1* | *IL12A, TRIM59* |
| rs10036748 | 5 | T/C | *TNIP1* | *TNIP1, ANXA6, GPX3* |
| rs2431697 | 5 | T/C | *MIR3142HG* | *PTTG1, SLU7* |
| rs57668933 | 13 | C/T | *ELF1* | *ELF1* |
| rs2069235 | 22 | A/G | *SYNGR1* | *PDGFB, MGAT3, RPL3* |

We examined these SNPs in detail. rs57668933 (intron of lymphoid cell transcription factor E74- like factor 1, *ELF1*), at LOC_125 (Chr 13), controls *ELF1* expression (**Figure 3a-b**). The protective T allele correlates with higher *ELF1* expression in T-cells, B-cells, and monocytes in healthy controls (**Supplementary Figure 7**) and shows high allele-specific chromatin accessibility (**Figure 3c**) and enhancer activity (**Figure 3d**). *ELF1* has been previously reported as an SLE risk gene (lead SNP rs7329174(47))–we show that rs57668933 is instead the likely causal SNP, with the risk allele yielding lower chromatin accessibility and *ELF1* expression. ELF1 represses FcRγ expression(48); SLE patients’ T-cells express essentially no ELF1 but high levels of FcRγ, which activates immune reactivity and promotes nephritis(49). ELF1 also regulates antibody heavy chain production in B-cells. This SNP disrupts universally conserved binding sites for the tumor suppressors p63 and p73 (**Supplementary Figure 6a**), both with strong immune contributions.

All six SNPs show much experimental evidence linking them to SLE (**Table 2, Figure 3, Supplementary Figure 8a-e**). Most disrupt highly conserved binding sites of critical immune transcription factors (**Supplementary Table 14, Supplementary Figure 3**). Target genes and disrupted transcription factors are known autoimmune risk genes, implicated in multiple diseases (**Supplementary Table 5**). Many selected SNPs are far from index SNPs and do not appear in the literature, highlighting the pipeline’s ability to localize signals in large GWAS peaks.

Beyond the six most highly selected SNPs (**Table 2**), our Tier1 hits and associated targets were very strongly enriched in immune-related genes. High-tier SNPs were also greatly enriched in SNPs flagged as deleterious by other methods. Overall, putative risk loci and target genes were overwhelmingly enriched in immune genes, with many being known risk for SLE, rheumatoid arthritis, systemic sclerosis, Crohn’s disease, Sjögren’s syndrome, primary biliary cholangitis, and particularly inflammatory bowel disease. We experimentally validated a high-priority SNP with CRISPR/Cas9 gene activation/silencing, confirming that this site indeed has dramatic enhancer activity, likely underlying SLE association. This experimental support for SNPs and loci prioritized by our analysis supports its utility in selecting likely underlying SNPs from GWAS peaks.

Our study provides valuable insights into the functional variants and target genes associated with SLE, but has two major limitations. Firstly, the sparse MPRA data utilized in our analysis may result in some loci having unflagged causal variants, potentially leading to missed associations. Secondly, the sparse caQTL data restricts the strongest conclusions to B-cells, limiting the generalizability of our findings to other cell types. To address these limitations, it is crucial to generate more MPRA data and caQTL data in diverse cell types. This would refine the existing loci, identify additional loci, and enhance the applicability of our pipeline to a broader range of diseases. Furthermore, future studies should focus on verifying causality and elucidating underlying biochemical mechanisms, utilizing our SLE dataset as a roadmap.

Another limitation of our study applies to all genetics projects: limited power to resolve rare variants. Various groups have shown that SLE risk loci, including our risk locus *BANK1*(50), are enriched in rare variants (sometimes strongly) associated with disease. Increasing sample sizes, and performing meta-analyses such as we do, increase power for resolving such associations – although follow-up candidate-gene experiments are required to analyze and validate rare variants. It is likely that some of our loci manifest at least somewhat through rare SNPs.

In conclusion, we demonstrate and validate a comprehensive analysis pipeline useful for diverse post-GWAS studies. We anticipate that this work will inspire future research to verify causal relationships and uncover the intricate biochemical mechanisms underlying SLE and related diseases. The SLE dataset we generated will serve as a roadmap for future studies verifying causality and establishing underlying biochemical mechanisms.

## Data Availability

All data produced in the present study are available upon reasonable request to the authors

